# Radiographic Imaging of Power Injectable Medical Access Ports as a Supplemental Identification Tool

**DOI:** 10.1101/2022.02.15.22270761

**Authors:** Nikhil Goyal, Huy M. Do, Alexandra P. Toscano, Lillian G. Spear, Lindsey A. Hazen, Les R. Folio

**Affiliations:** Perelman School of Medicine, University of Pennsylvania, Philadelphia, PA, USA; UT Health Science Center in Houston at McGovern Medical School, Houston, TX, USA; University of Maryland, College Park, MD, USA; Center for Interventional Oncology, Clinical Center, National Institutes of Health, Bethesda, MD, USA; Moffitt Cancer Center, Tampa, FL, USA

**Keywords:** Power injection, medical access ports, X-ray, Computed tomography, dual-energy subtraction radiography

## Abstract

**Purpose:** Our aims were to describe characteristic radiographic features of two power injectable medical access ports (MAPs) on various imaging modalities for rapid and precise identification; and to demonstrate the value of this approach in identifying other types of MAPs via “pictorial atlas”.

**Methods:** We analyzed two commonly seen MAPs at our clinical center, Smart Port® CT-Injectable Port and PowerPort® M.R.I.® Implantable Port. Photographs of these two MAPs were retrospectively compared with identity-verified MAPs seen on chest X-ray (CXR), computed tomography (CT) and dual energy subtraction radiography (DESR) images from routine patient encounters at our clinical center. Visualized radiographic features were used for MAP differentiation and identification.

**Results:** Based on selected patient case examples for these two MAPs, physical characteristics seen on imaging were used for MAP identification. These properties included port body and chamber shape; location and number of suture holes; and radiopaque and radiolucent features. Each imaging modality provided a unique set of radiographic features and highlighted specific components of each MAP for rapid and precise identification. CXR offered better visualization of unique MAP features compared to CT.

**Conclusions:** Radiographic imaging can serve as a tool for medical staff to quickly identify MAPs. Hospital-specific “pictorial atlases” can be developed to display MAPs along with their associated distinctive radiographic and physical features for rapid and precise identification. This may be useful for large referral centers that see a wide array of MAPs by mitigating complications associated with MAP misidentification and usage, thus improving patient care.

## Introduction

Power injectable intravenous (IV) medical access ports (MAPs) are surgically implanted indwelling devices used for frequent administration of therapeutic agents and contrast-enhanced diagnostic imaging exams, such as computed tomography (CT)^1^ or magnetic resonance imaging (MRI). There are numerous commercially available MAPs from different vendors, each with their own specific usage parameters (e.g. proprietary needles, injection pressures and flow rates).

Since MAPs may or may not be power injectable with differences in usage and product specifications, it is essential to be able to consistently verify the identities of these indwelling devices. Current methods of MAP identification include product identification cards or bracelets, electronic medical record (EMR) documentation, and palpable points or “bumps” for certain types of power injectable MAPs.^2–4^ When MAP identities cannot be verified using these methods, hospital staff often resort to peripheral IV placement for power injection to avoid complications associated with port misidentification and misuse, potentially causing further delays in diagnostic and therapeutic interventions.^5^

Port misidentification and power injection at inappropriate pressure and flow rate settings can cause catheter rupture, leading to contrast extravasation and catheter fragment embolism that may require surgery and eventual port replacement.^3,6^ One possible complication resulting from delayed recognition of catheter fragmentation is fibrin sheath formation related to thrombus development around the catheter tip, which enhances difficulty of extraction due to vessel wall adherence and leads to infections via biofilm formation.^7^ Notably, catheter migration, fibrin sheath formation and bloodstream infection accounted for 1.3%, 0.61% and 5.11% of all total port catheter-related complications assessed in a large retrospective study, respectively, which are important factors that endorse the need for precise port identification.^8^

Most chest MAPs have centrally located radiopaque or radiolucent “CT” lettering, which is indicative of their capability to handle power injection and is helpful for more precise MAP identification and categorization on chest X-ray (CXR) and CT.^2,4,9,10^ Furthermore, since MAPs are composed of multiple different materials, MAP identification via radiographic imaging could be further elucidated using dual energy subtraction radiography (DESR). DESR exploits the photon energy-dependent attenuation coefficient of various materials, allowing for image acquisition at both low and high energies to improve material characterization.^11,12^

In this paper, we describe characteristic radiographic features of two commonly seen power injectable MAPs identified at our clinical center on CXR, CT and DESR imaging through representative examples. The two power injectable MAPs selected in this study are important to differentiate and identify since they each require their own vendor-specific non-coring Huber needles for access. We illustrate the potential value of radiographic imaging for rapid and precise MAP identification based on their unique characteristics and features. In addition, we provide a review of imaging parameters and techniques to optimize MAP viewing. Furthermore, we demonstrate how our approach can be extrapolated to identify other types of MAPs and improve patient care, especially at large referral centers that frequently encounter numerous types of MAPs.

## Methods

The NIH Clinical Center Institutional Review Board reviewed this project and waived ethical approval because this project qualified as a quality improvement initiative. Data collected was anonymized, not identifiable, and did not require informed consent.

In this study, we analyzed two MAPs: the Smart Port® CT-Injectable Port (AngioDynamics, Latham, NY) and the PowerPort® M.R.I.® Implantable Port (Bard Access Systems, Salt Lake City, UT). We will refer to these as “Smart Port” and “PowerPort,” respectively. These two MAPs were selected for evaluation because each required a different vendor-specific non-coring needle for access, highlighting the importance of fast and precise MAP identification in preventing errors and mitigating delays in patient care. Various components and structural features of MAPs are shown in Figure 1.

**Figure 1:**
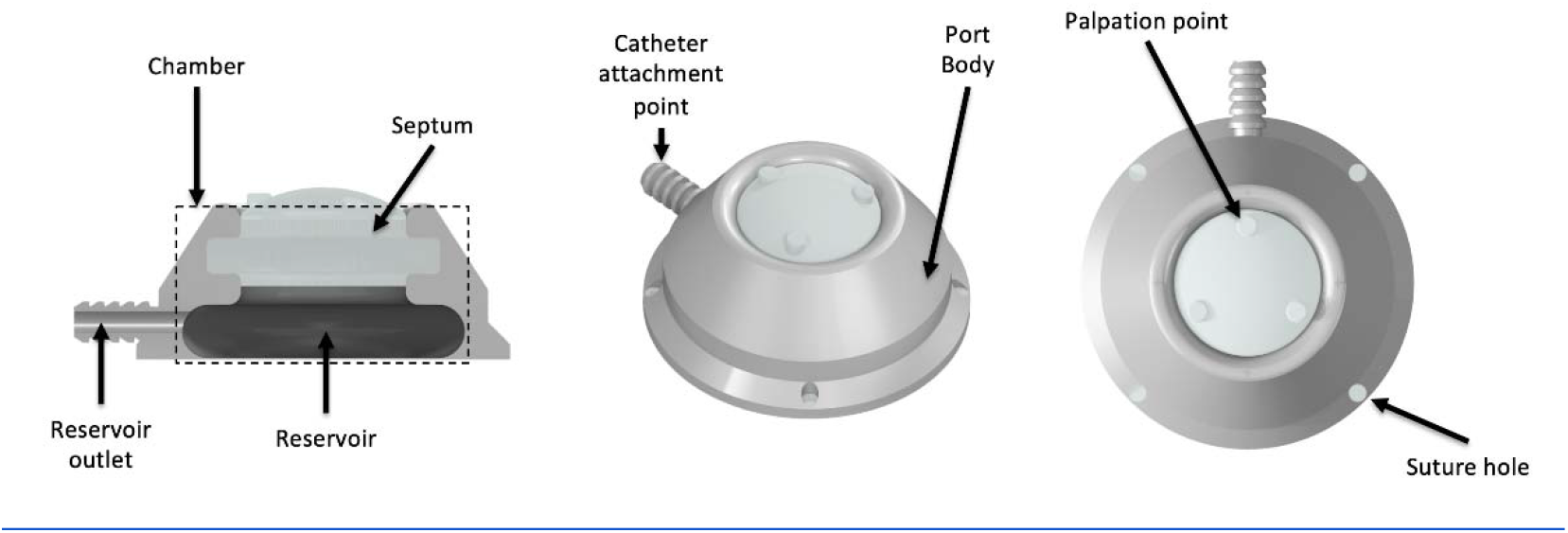
3D model of a generic medical access port (MAP) created by N.G. using Autodesk Inventor Professional 2021. This model was created to illustrate the basic features of MAPs and was not based on any particular commercially-available MAP. The MAP “chamber” is the interior cutout portion of the port body, which contains the septum and reservoir.

Photographs of the *Smart Port* and *PowerPort* were taken by authors L.H. and H.D. using available device samples at our clinical center. Photographs were compared with corresponding radiographic images acquired from our clinical center, providing qualitative information about the physical characteristics of these ports.

We retrospectively reviewed dual-energy subtraction PA or AP (less common) CXR exams that were acquired using GE Healthcare Discovery XR656 (Milwaukee, WI), a dual-exposure system capturing two images (one at 60 kV, one at 120 kV) with a 200 ms delay and effective mAs of 3-6. By convention, the standard CXR image was acquired at 120 kV and the bone-selective image was acquired at 60 kV. A post-processing algorithm then subtracted highly attenuating components and substances (e.g. bone, metal) from the standard CXR image to produce a soft tissue-selective image. For each exam, approximately 1-4 images were pushed to PACS with the inclusion of an accompanying lateral view. The standard CXR, bone-selective and soft tissue-selective DESR images selected as representative examples of both the *Smart Port* and *PowerPort* originated from image series acquired on the GE Healthcare Discovery XR656.

CT exams were obtained on Siemens Somatom Force (Erlangan, Germany) at 120 kV with effective mAs of 150 (dose modulated with ADMIRE version 2-4 MBIR). Images pushed to PACS included 2×1 overlapping soft and sharp algorithms for an average of 1500 images per study. A majority of CT CAP exams were completed with only IV contrast in venous phase and water or oral omnipaque for PO contrast.

For the purposes of this study, all radiographic images were taken from patient cases at our clinical center and carefully cropped to show only MAPs while excluding surrounding pathology since cases were selected solely to demonstrate MAP properties on imaging. To obtain off-axis reconstructions of these MAPs on CT, images were manually repositioned using the multiplanar reformat (MPR) and reference tools in our PACS (VuePACS Philips, Amsterdam, Netherlands), allowing for more direct comparison with corresponding CXR images. The multiplanar volume reconstruction (MPVR) feature in our PACS was used to generate 3D images of the MAPs.

Radiographic images from cases at our clinical center were acquired as part of clinical workups. Therefore, acquisition parameters for the discussed imaging modalities were not modified to optimize viewing of MAPs. For example, CXR and CT exams were not specifically ordered for the sole purpose of evaluating MAPs, but rather for baseline or follow-up evaluation of disease status in clinical trial patients at our clinical center. In this paper, we describe how routine patient imaging can be exploited to characterize and differentiate between two different types of MAPs.

## Results

Representative images of the *Smart Port* and *PowerPort* from selected radiology studies at our center are shown in Figures 2 and 3, respectively.

**Figure 2:**
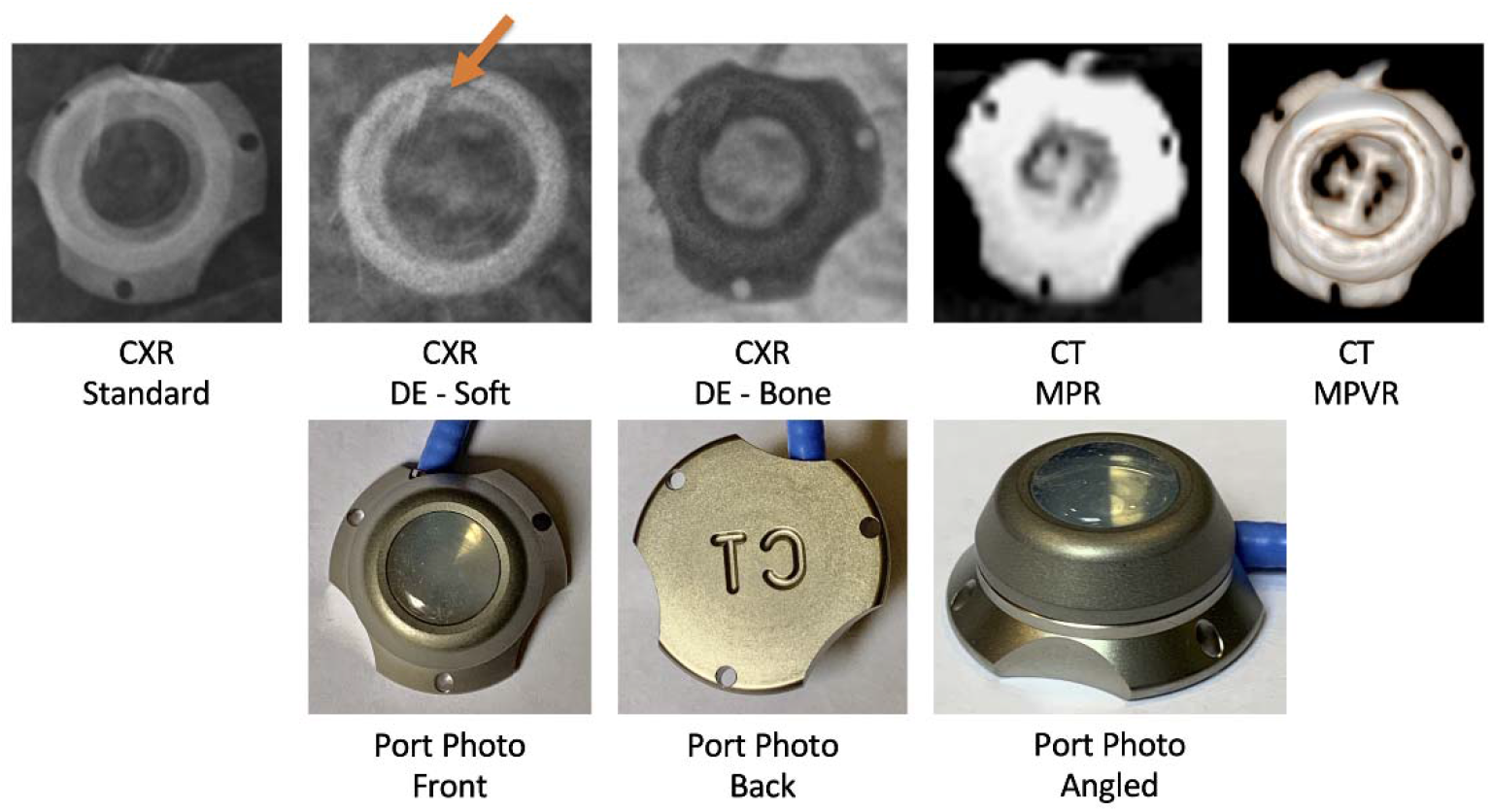
(Top row) Radiographic images of the Smart Port® CT-Injectable Port (AngioDynamics) acquired from clinical scans of a patient at our institution demonstrating the radiolucent “CT” letters overlying the center port chamber. Orange arrow in CXR DE - Soft image indicates the visible tangential reservoir outlet, which is unique to this MAP. (Bottom row) Photographs of the MAP sample obtained from our institution *(Photo credit to author L.H. and H.D.)*.

**Figure 3:**
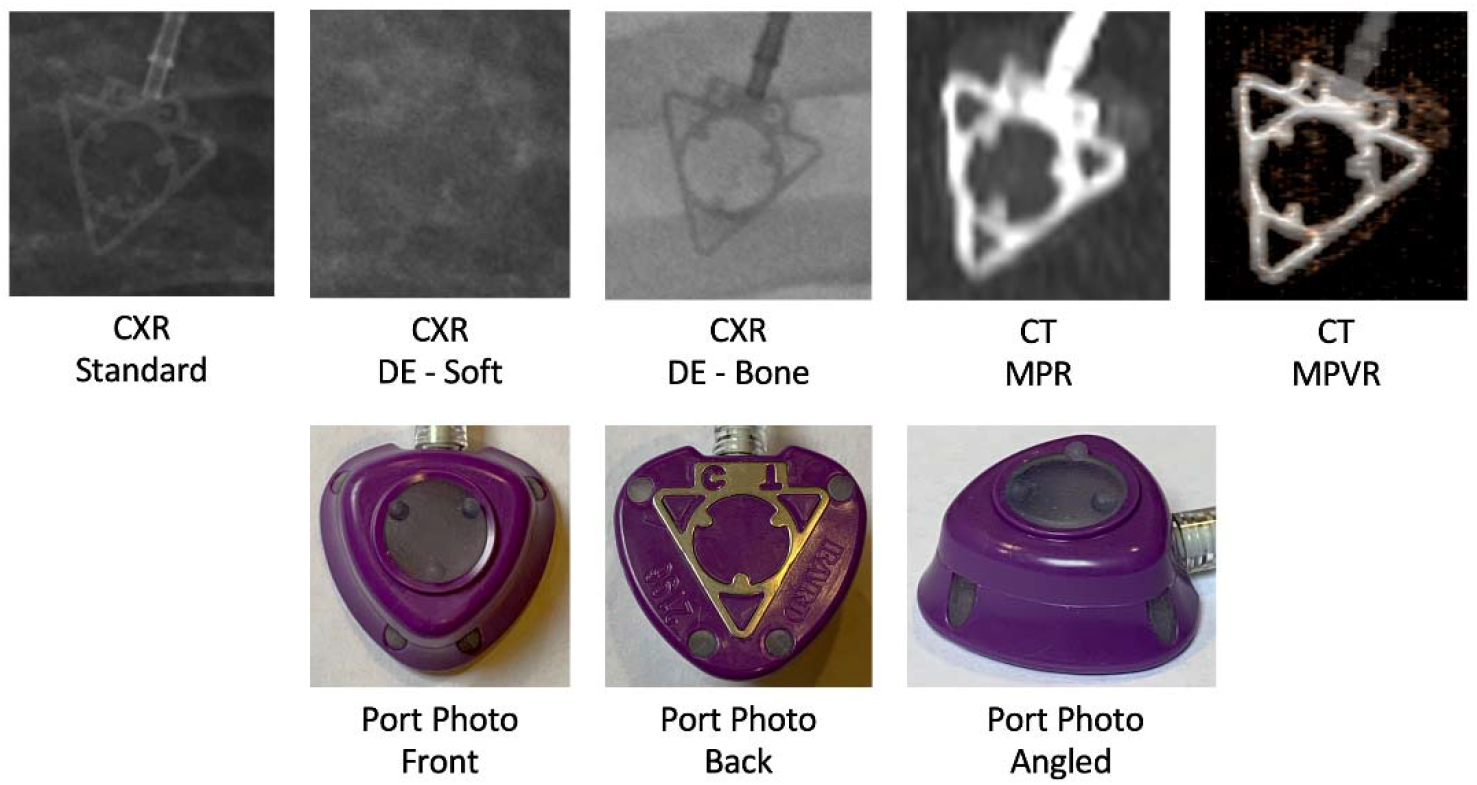
(Top row) Radiographic images of the PowerPort® M.R.I.® Implantable Port (Bard Access Systems) acquired from clinical scans of a patient at our institution demonstrating the radiolucent “CT” letters outside the port chamber housed within the characteristic metallic triangular shape on the bottom surface of the port. (Bottom row) Photographs of the MAP sample obtained from our institution *(Photo credit to author H.D.)*.

### Smart Port ® CT Power-Injectable Port (AngioDynamics)

Figure 2 is from a male in his 30s with a clinical history of cancer with lung metastases who has a *Smart Port*. From the standard CXR image in this example, the port body has a unique shape and contour that is easily identifiable upon initial observation. As a metal, titanium more greatly attenuates X-rays and is more dense than the cut-out suture holes^13^, which allows for easier recognition and distinction of the suture holes themselves relative to the titanium port body on standard CXR.

Another identifying feature of this port is the radiolucent “CT” letters engraved on the bottom surface of its body. These letters appear radiolucent on the standard CXR image, which is helpful in successfully identifying this port and its capability to support power injection. This feature is depicted in Figure 2 for both standard CXR and CT images.

The round chamber of the *Smart Port* appears more radiopaque than its titanium body on standard CXR, which suggests that the material composition of the port chamber could be made of a more radiopaque element or polymer than titanium, or the radiographic appearance could be attributed to the unique elliptical geometry of the chamber itself.^9^ Additional information can be determined from the DESR images for further verification as needed.

The bone-selective DESR image is acquired at 60 kV on our dual-exposure system, exploiting the effect in which calcium-containing structures (e.g. bone) have higher attenuation coefficients at lower photon energies than soft tissue.^12^ Since titanium has an attenuation coefficient similar to that of calcium, the titanium body of this port appears radiopaque in the bone-selective DESR image. Notably, a weighted subtraction removes highly attenuating elements of the bone-specific image from the standard image, producing a soft tissue-selective DESR image via post-processing in which only the round chamber of the port is visible (shown in Figure 2, “CXR DE - Soft” image).^11,12^ Another subtle finding on this image is the radiolucent reservoir outlet seen within the round chamber (shown in Figure 2, arrow in “DE CXR - Soft” image), which corresponds to the tangential outlet that leads to the catheter and is unique to the design of this port.^9^ Furthermore, the port appears to be slightly angled in the soft tissue-selective DESR image, which allows us to visualize two nearly superimposed circular structures that are radiopaque and correlates with the elliptical-shaped chamber design of this port.

The off-axis MPR CT image of this port illustrates the engraved “CT” letters more clearly than the DESR images due to the 3D acquisition of CT images relative to the 2D projection of CXR images. The overlying silicone port septum and underlying pulmonary vasculature may obscure visualization of the engraved “CT” letters on CXR since these images are captured as 2D projections. Alternatively, CT images can be manually reconstructed in MPR in PACS to reorient the port for optimal off-axis visualization of the radiolucent “CT” letters. Figure 2 also shows this port in MPVR mode, another form of CT image reconstruction possible on our PACS. MPVR is most useful for visualizing the shape and contour of the port body, the engraved “CT” letters and the three suture holes, all of which are helpful for successful identification of this port.

### PowerPort ® M.R.I. ® Implantable Port (Bard Access Systems)

Figure 3 is from a male in his 30s with a clinical history of lymphoma who has a *PowerPort*. From the standard CXR image in this example, the triangular titanium radiopaque identifier on the bottom surface of this port is clearly visible along with the radiolucent “CT” lettering to indicate the port’s capability to support power injection and allows for quick recognition of proper port orientation. The plastic port body itself (shown in Figure 3, purple component seen in port photos) is not visible on the standard CXR image.

The bone-selective DESR image was acquired at 60 kV on our dual-exposure system, and provides a similar level of detail about port structure and identity as the standard CXR image with clear visualization of this port’s unique triangular-shaped titanium radiopaque identifier^10^. The soft tissue-selective DESR image generated via post-processing removes the titanium component of the *PowerPort*, but is not helpful for identification. The plastic port body does not appear on the standard CXR or DESR images because the attenuation coefficient of plastic may be similar to that of soft tissue as depicted in Figure 3. Since CXR images are acquired as 2D projections, normal pulmonary vasculature and airways in addition to underlying pathology (if present) can obscure visualization of the plastic port body, especially in the soft tissue-selective DESR images.

Similar to the standard CXR and bone-selective DESR images, the off-axis MPR and MPVR CT images of the *PowerPort* clearly show its underlying titanium structure, but the “CT” lettering is more obscured whereas the radiopaque titanium identifier is prominent. Additionally, there is a low density region surrounding the “CT” lettering and port catheter on the off-axis MPR and MPVR CT images as seen in Figure 3, which is likely a portion of the plastic port body but cannot be concluded due to incomplete visualization.

## Discussion & Conclusion

Based on the representative case examples we have selected for two commonly encountered power injectable MAPs at our clinical center, there are numerous physical characteristics of MAPs that can be differentiated on radiographic imaging and used for identification as shown in Figures 2 and 3. These properties include the overall shape of the port body and port chamber; location and number of suture holes; and presence of radiopaque and radiolucent features. Together, these characteristics provide a unique set of radiographic features for these two power injectable MAPs on standard CXR, DESR and CT, which can be used for rapid and precise MAP identification.

In this study, we find that each imaging modality provides useful information for identifying both the *Smart Port* and *PowerPort* by highlighting specific components of these MAPs. We believe CXR is not only sufficient, but offers better visualization of unique MAP features compared to CT. Proposed guidelines for using various imaging modalities to visualize features of MAPs are summarized in Table 1. Routinely acquired for many patients at our clinical center to evaluate disease status, the standard CXR displays many qualitative port features that are helpful for identification. For the *Smart Port*, the standard CXR image clearly illustrates the radiopaque titanium port body with three suture holes, the unique round chamber and the radiolucent “CT” letters engraved on the bottom surface of the port. The plastic port body of the *PowerPort* is not visible on the standard CXR image, but the radiopaque titanium identifier with radiolucent “CT” letters on the bottom surface is unique and easily recognizable, which allows for quick identification of this port and its capability to handle power injection.

**Table 1:**
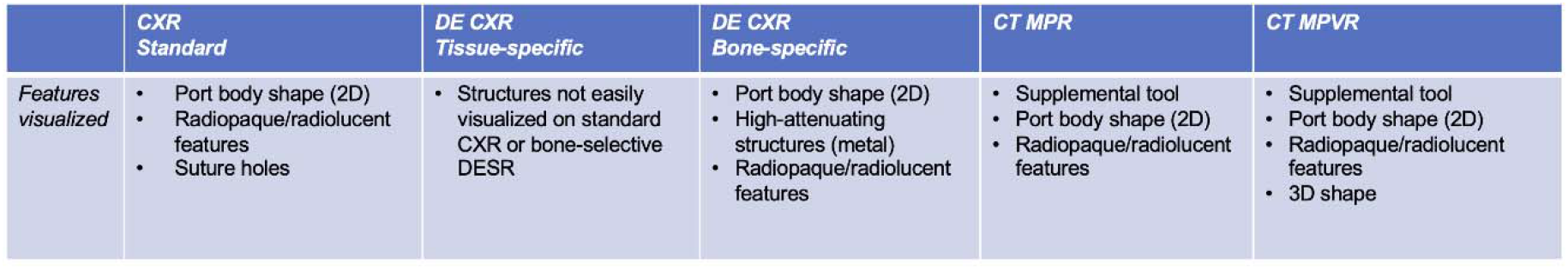
Basic guidelines for using different imaging modalities to identify unique features of MAPs.

DESR exploits the concept that different materials have different attenuations at high and low photon energies.^11,12^ Acquired at 60 kV, the bone-selective DESR images are most suitable for visualizing materials and structures that are highly attenuating at lower photon energies, such as calcium and titanium.^11,12^ Since both the *Smart Port* and *PowerPort* have unique identifiers composed of titanium, the bone-selective DESR images provide optimal visualization of these features as depicted in Figures 2 and 3. Generated via post-processing, the soft tissue-selective DESR images are typically used to better visualize lung parenchyma and soft tissue without being obscured by bone or other highly attenuating materials.^12^ For MAPs, the soft tissue-selective DESR images are most useful in identifying structures that are not easily recognizable on the standard or bone-selective CXR images. This is particularly true for the tissue-selective DESR image of the *Smart Port* (Figure 2) in which we only see a structure that resembles the round chamber while most of the titanium body has been removed by post-processing. This radiopaque chamber on the tissue-selective DESR image could be used as a unique identifier for the *Smart Port*.

As a standalone imaging modality, CT is suboptimal for viewing and identifying MAPs because its images are acquired in 3D, typically at 2 mm and 5 mm slice thicknesses on our CT scanner. Furthermore, since CT images need to be manually reoriented off-axis on PACS in MPR mode to achieve more complete visualization of these MAPs, the reconstructed CT images do not delineate port features as well as CXR images. When available, a recent CT is advantageous as a supplemental MAP identification tool to CXR and may showcase certain features that are more difficult to visualize on CXR. In our two examples, the engraved cutout “CT” letters on the *Smart Port* and the radiopaque titanium indicator on the *PowerPort* are easily seen on CT images. Notably, the CT MPVR images are useful for determining the 3D shape and geometry of MAPs. This is seen in our example of the *Smart Port* in which the portion of the port body surrounding the round chamber appears raised relative to the bottom part of the body. To a lesser extent, the plastic port body of the *PowerPort* is partially visualized in the CT MPVR image.

Figure 4 is a detailed graphic containing labeled features that can be used to identify the *Smart Port* and *PowerPort* on standard CXR, bone-selective and soft tissue-selective DESR, and CT images. This graphic provides salient examples that further illustrate the concepts described in Table 1.

**Figure 4:**
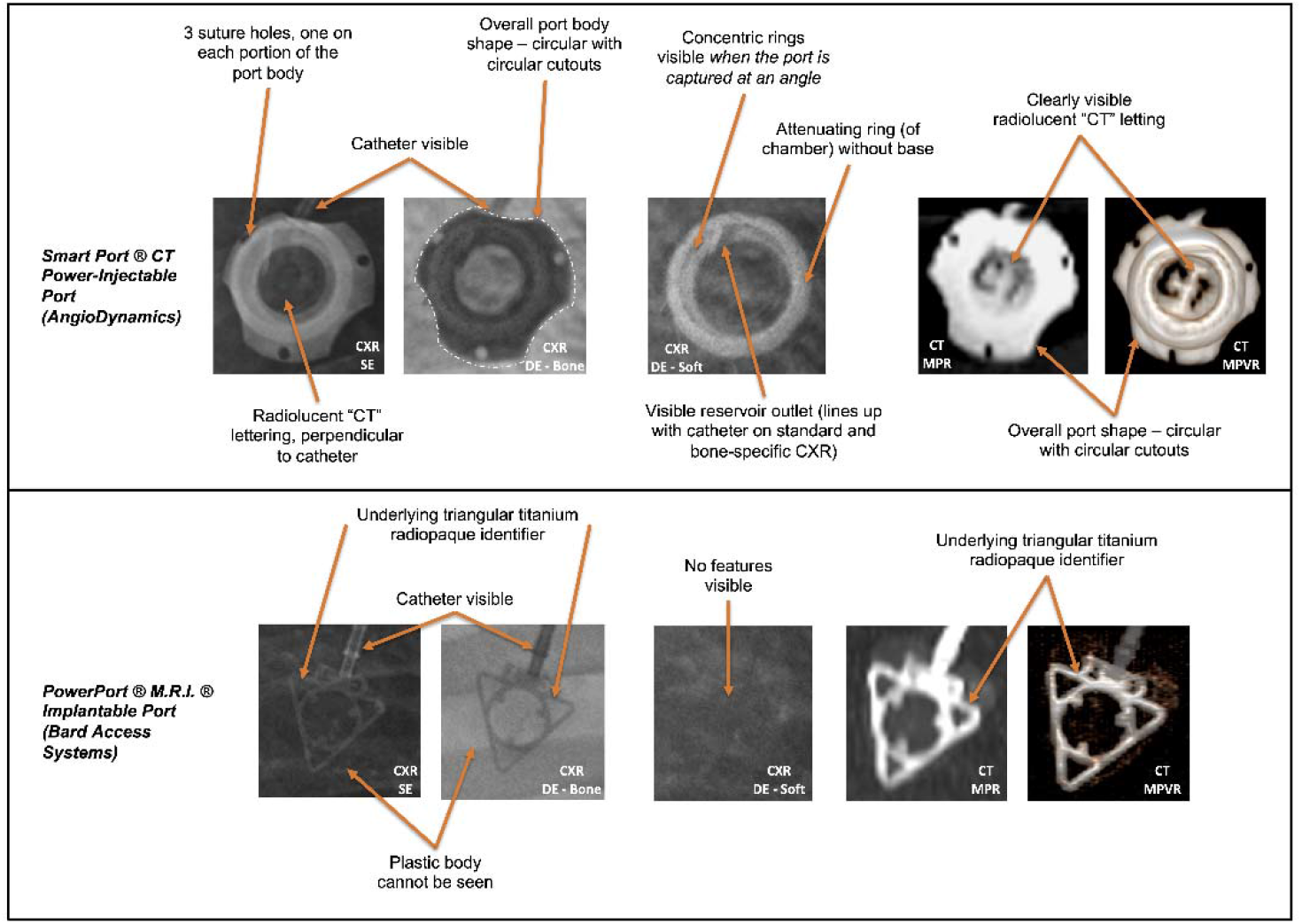
Important MAP features on radiographic imaging allowing for identification of the Smart Port® CT-Injectable Port and PowerPort® M.R.I.® Implantable Port.

By utilizing the guidelines in Table 1 to generate unique identification features (and unique combinations of features) for a number of different MAPs, an institution-specific MAP identification “pictorial atlas” can be created to show photographs and radiographic images of MAPs along with key identification features for use in clinical practice.^14^ Furthermore, MAP specifications such as flow rate, maximum pressure, and needle size-flow compatibility could be included in the pictorial atlas to assist clinicians and hospital staff by ensuring proper port usage and mitigating errors associated with port misidentification. Figure 5 depicts an example pictorial atlas consisting of representative CXR images from eight commonly seen power injectable MAPs at our institution along with their vendor-specific names and specifications. Additionally, the identities of the two MAPs evaluated in this study and displayed on this atlas, Smart Port® CT-Injectable Port and PowerPort® M.R.I.® Implantable Port (shown in Figure 5, orange dashed line), was verified by electronic medical record (EMR) documentation. The identities of the other six MAPs seen in Figure 5 were unconfirmed but suggested based on port design and qualitative comparison with corresponding port photographs.

**Figure 5:**
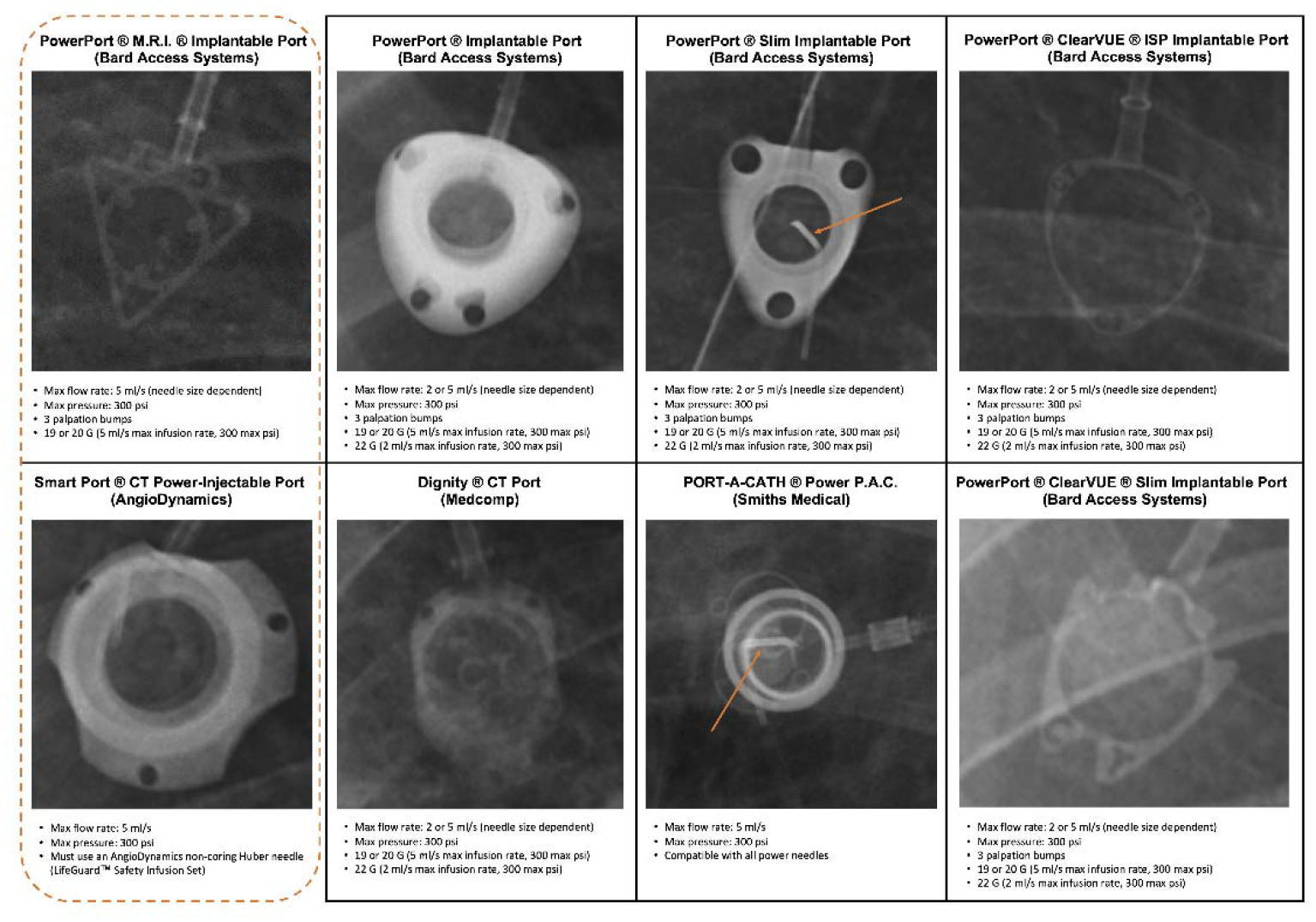
Chest X-ray (CXR)-based pictorial atlas of eight power injectable medical access ports (MAPs) commonly encountered at our institution, a large U.S. referral center for clinical trial cancer patients. All eight MAP CXR images are labeled with their vendor-designated names and specifications, including maximum injection flow rates and pressures, and needle size-flow rate compatibility. The supplemental MAP information in this atlas may not be fully inclusive but can be modified and customized based on institution-specific needs to facilitate fast and precise MAP identification for clinical use. Note that the selected CXR images for PowerPort ® Slim Implantable Port and PORT-A-CATH ® Power P.A.C. on this atlas depict each MAP being accessed by a non-coring Huber needle (orange arrows), which is not a component of the MAPs themselves.

There are several limitations to the MAP identification techniques used in this study. Firstly, there are a variety of X-ray and CT scanners used by different institutions and practices, which can affect the generalizability of our approach to identify power injectable MAPs using imaging. Numerous factors can impact the final CXR and CT images that are produced, including image acquisition parameters and post-processing algorithms. All CXR images used as representative examples of the *Smart Port* and *PowerPort* were acquired on the dual-exposure GE Healthcare Discovery XR656, which allows for a higher signal to noise (SNR) ratio and, hence, improved image quality. However, there is a 200 ms delay between the acquisition of the standard CXR at 120 kV and the bone-selective image at 60 kV, possibly leading to misregistration artifacts in the soft tissue-selective image (generated via post-processing weighted subtraction) due to normal physiologic processes such as cardiac and respiratory motion.^12^ Additionally, all CT images of the *Smart Port* and *PowerPort* were acquired on the Siemens Somatom Force, which applies a model-based iterative reconstruction (MBIR) algorithm known as advanced modeled iterative reconstruction (ADMIRE) to reduce noise in both the raw data and image itself to improve image quality and facilitate dose reduction.^15^ This can lead to smoothing effects of certain anatomical features and change their appearances on imaging depending on the applied ADMIRE strength.^15^ However, the representative CXR and CT image examples used in this study were acquired as part of our patients’ routine clinical workups, demonstrating that image protocols, acquisition parameters and post-processing algorithms do not necessarily need to be optimized in order to view and identify unique MAP features. Furthermore, our approach is likely applicable and generalizable to other referral centers and practices regardless of the type of CXR and CT scanner used.

Another limitation is the differing capabilities of various PACS at other institutions, which may affect the generalizability of our findings. The PACS at our clinical center offers a wide range of tools, including MPR and MPVR modes to manually manipulate CT images “on the fly”. In conjunction with the reference tool, these modes allowed us to view our two case examples in off-axis planes and in 3D to provide more information about specific MAP features. While PACS at other institutions may not offer the same functionalities, there may be other unique tools that can be used to optimize viewing MAPs on CT.

MAP angle and location can also adversely impact port visualization. This is particularly true for CXR because these images are acquired as 2D projections and cannot be manipulated on PACS in the same way as CT data. Underlying anatomical structures (e.g. bone, lung parenchyma, vessels, airways) can also obscure MAP viewing on CXR images. To improve visualization, additional imaging modalities like DESR and CT can be exploited to highlight specific identifying regions and features in MAPs. Further details are provided in Table 1.

As this is an initial exploration into the potential use of imaging for MAP identification, this is not a fully inclusive study of all available MAPs. Further research is necessary to validate this MAP identification approach using a larger sample size of commercially available MAPs with images to determine if unique radiographic features for each MAP are consistent across different X-ray and CT scanners. Accuracy and inter-reader concordance testing are also needed to determine whether this identification approach would be appropriate for use in a clinical setting. Once validated, artificial intelligence (AI) can be explored as an option to automate MAP identification in an augmented clinical workflow, assisting radiologists by quickly and precisely identifying MAPs seen on imaging with further categorization by vendor and type.^16^

## Supporting information

Take-Home Points

## Data Availability

All data produced in the present study are available upon reasonable request to the authors.

## Abbreviations

MAP: Medical Access Port
CXR: Chest X-ray
CT: Computed Tomography
IV: Intravenous
EMR: Electronic Medical Record
DESR: Dual Energy Subtraction Radiography
MPR: Multiplanar Reformat
MPVR: Multiplanar Volume Rendering
PACS: Picture Archiving and Communication System
MBIR: Model-Based Iterative Reconstruction
ADMIRE: Advanced Modeled Iterative Reconstruction
AI: Artificial Intelligence

## Acknowledgments

This is a HIPAA-compliant prospective IRB-exempt quality improvement initiative. We would like to thank Michael Spivey for his participation and contributions to this work. We would also like to thank Alicia A. Livinski, NIH Library Editing Service, for reviewing the manuscript. Supported in part by the Intramural Research Program of the National Institutes of Health Clinical Center. The content of this manuscript is solely the responsibility of the authors and does not necessarily represent the official views of the National Institutes of Health.

