## Supplementary material for "Radiographic Imaging of Power Injectable Medical Access Ports as a Supplemental Identification Tool": Take-Home Points

- Radiographic imaging can serve as a tool for radiologists, radiologic technologists and nursing to quickly identify MAPs by exploiting CXR and CT images to visualize distinctive MAP features.
- Taking advantage of physical appearances and material compositions, CXR and CT imaging can be used to generate unique sets of features to identify MAPs.
- An institution can create its own “pictorial atlas” that allows its clinicians and medical staff to distinguish between MAPs that they commonly encounter.
- This proposed MAP identification technique could prove useful for large referral centers where patients do not initially have their MAPs placed and helps mitigate complications associated with MAP misidentification and usage, thus improving patient care.
